# Age-related changes of the brain’s arterial network assessed with machine learning segmentation

**DOI:** 10.1101/2025.09.10.25335518

**Authors:** Jorge T. Gomez, Rachit Saluja, Mert R. Sabuncu, Henning U. Voss

## Abstract

**Purpose:** To provide a tool for the automatic segmentation of an arteriogram of the brain from MRA images and the estimation of arterial tortuosity as a summary marker.

**Methods:** A deep learning model was trained and validated on a previously published set of semi-automatically segmented brain arteriograms. We tested whether arterial tortuosity estimated from a large number of age-representative subjects (N = 478) would reproduce previously published statistics of increasing tortuosity with age.

**Results:** The tool provides a segmentation of the arteriograms from MRA images of varying resolution and quality. The arterial tortuosity estimated from the automatically segmented brain arteriograms approximately matched their previously published statistics. Further, a highly significant increase of tortuosity with age was observed in the large dataset with 478 subjects (p = 9 ×10^-8^).

**Discussion and conclusion:** The proposed ASN (Angiogram Segmentation Network) algorithm can provide the radiologist with a clean arteriogram of the brain, without the need for manual segmentation by the MR operator. Moreover, it offers arterial tortuosity as an instant quantitative metric that can augment the qualitative visual reading of the MRA.

## 1. Introduction

Tortuosity, defined as the degree of curving, curling, looping, winding, or kinking of blood vessels^1, 2^, is increasingly recognized as a morphological hallmark of cerebrovascular aging^3^. Both histopathological and imaging studies have shown that the prevalence and severity of vascular tortuosity begin to increase in middle age and continue to progress in older adults^4-10^. Similar age-related changes in arterial tortuosity have also been observed in animal models of aging^11, 12^. Moreover, increased venular tortuosity has been associated with early markers of cerebral small vessel disease and cognitive decline, suggesting a potential role in the pathogenesis of vascular cognitive impairment^13^. Accurately quantifying vascular tortuosity is, therefore, providing important insights into the biological aging of the brain and its related pathologies^1, 14-19^. For instance, tortuous vessels in the white matter are frequently surrounded by perivascular cavities, indicative of parenchymal tissue loss. Additionally, the increased length and curvature of these vessels can lead to reduced kinetic energy and perfusion pressure, thereby increasing the risk of ischemic damage^20^. The mechanical buckling hypothesis^1, 21^ proposes that tortuosity arises as a result of vessel buckling due to elevated luminal pressure or weakened surrounding tissue. This model provides a theoretical link between vascular tortuosity and arterial pulsatility^22-28^, although further research is necessary to establish the nature of this relationship.

Magnetic resonance angiography (MRA) offers a non-invasive modality to visualize and assess cerebral vasculature in vivo. The most widely used technique, time-of-flight (TOF) MRA, enables high-resolution volume imaging of intracranial arteries at the level of the macrocirculation^29^ in the main arteries and veins^30-33^. It relies on the signal enhancement of inflowing blood and the simultaneous suppression of stationary signals. Time-of-flight MRA can be tailored to emphasize either arterial or venous structures. When applied to highlight arteries, the technique relies on the inflow of unsaturated blood entering the imaging volume via the carotid and basilar arteries in the neck, producing what is commonly referred to as an arteriogram. The arteriogram is the focus of the present study. Conversely, when TOF-MRA is configured to enhance venous signal, it visualizes blood draining from the superior regions of the brain, resulting in a venogram.

While TOF-MRA is capable of producing arteriograms with spatial resolution in the millimeter range or slightly higher, the resulting images are not pure representations of the arterial vasculature. Residual signals from stationary brain parenchyma, extra-cerebral tissues, and venous structures can contaminate the image and obscure arterial detail. In clinical settings, it is common practice for the MRI operator to manually remove these unwanted signals using editing tools on maximum intensity projections (MIPs). However, this manual intervention is time-consuming and subject to inter-operator variability. Automated methods for generating clean arteriograms would not only reduce workload but also improve reproducibility by minimizing dependence on user interaction.

In addition, accurate segmentation of cerebral arteries from MRA images is a prerequisite for reliable tortuosity analysis. Traditional segmentation methods often struggle with the complex geometry and variability of cerebral vasculature. However, the advent of deep learning has revolutionized this domain^34^. Convolutional Neural Networks (CNNs), particularly architectures like U-Net^35^ and its variants, have demonstrated superior performance in segmenting intricate vascular structures. For example, studies have shown that 3D U-Net models can effectively delineate cerebral vessels, achieving high dice similarity coefficients and enabling the extraction of detailed vascular features^36^. These advancements not only enhance the accuracy of vessel segmentation but also streamline the process, making large-scale analyses of vascular tortuosity feasible.

Recent methodologies have employed advanced image processing techniques to extract vessel centerlines and compute tortuosity metrics. For instance, the use of skeletonization algorithms combined with path-finding methods allows for the quantification of vessel curvature and length, facilitating the assessment of age-related vascular changes^37, 38^. Such quantitative analyses are crucial for establishing correlations between vascular morphology and chronological aging.

In this contribution, we provide a tool that leverages an off-the-shelf neural network trained on publicly available segmented MRA data to fully automate cerebral artery segmentation and the estimation of tortuosity. Segmentation is performed for the full field-of-view covered in the MRAs and without manual selection of arterial seed points^7, 8^. Our study verifies the increase of tortuosity with age in a large sample of MRAs. The automatically estimated tortuosity provides a quantitative summary marker that could be used as an assessment of brain health and as a parameter in further correlation studies that characterize the biological brain age of a subject.

## 2. Materials and methods

### 1) Data

We use two main data sets, a training and an analysis data set.

Training dataset: We trained the ML model using the COSTA data set^39^. COSTA is a large (N=423), multi-center, semi-automatically annotated dataset of 3D TOF-MRA brain images, designed for cerebrovascular segmentation research. To create accurate cerebrovascular segmentations for the COSTA dataset, which are used as a gold standard here, the authors employed a semi-automated annotation pipeline combining deep learning with expert manual correction. The annotation process and review process required approximately 2 to 2.5 hours per volume, underscoring the labor-intensive nature of high-fidelity vascular segmentation.

Analysis dataset: To assess the correlation between age and vascular tortuosity, we combined two datasets with publicly available age metadata. The IXI dataset (https://brain-development.org/ixi-dataset/) consists of 570 MRA scans, and the BRAVA (http://cng.gmu.edu/brava) dataset includes 56 MRA scans. To prevent data leakage and to ensure that the tortuosity analysis reflects model performance on unseen data, 136 IXI scans that were part of the COSTA training set were excluded from the analysis. This ensures that the resulting tortuosity metrics are applicable for downstream clinical use. Finally, 12 scans had to be removed from the analysis as their age information was missing.

In total, the final analysis included 478 unique subjects, with ages ranging from 19 to 86 years. Imaging resolution varied by dataset: IXI-Guys and IXI-HH had a resolution of 0.47 × 0.47 × 0.8 mm, IXI-IOP had a resolution of 0.26 × 0.26 × 0.8 mm, and BRAVA had a resolution of 0.62 × 0.62 × 0.62 mm. From a visual inspection of the data MIPs, IXI-IOP had a superior quality in terms of detail but also in terms of better background signal suppression.

### 2) Data preprocessing

1. Intensity matching: MRA images often exhibit varying intensity values for the same tissue, largely due to differences in individual scanners, vendors, and imaging protocols. To address this, we applied z-score normalization^40^ or histogram matching to reduce intensity variability across datasets^39^.
2. Skull stripping: To improve the performance of machine learning models during training and inference, it is common practice to remove non-brain tissues such as the skull in brain MRI scans. The skull typically contains no relevant information for most neuroimaging tasks, including the one in this work. For this purpose, we used HD-Bet, a deep learning-based brain extraction tool that provides fast and accurate skull tripping across a variety of MRI modalities and scanner types^41^.

### 3) Training

Using the COSTA dataset, we utilized the nnU-Net framework^40^ to train a U-Net model. The resulting ASN (Angiogram Segmentation Network) model is an ensemble comprising five distinct models, each trained on a separate fold of the dataset using an 80/20 training-validation split. Each fold applied a series of data augmentation techniques, including Gaussian blurring, Gaussian noise addition, brightness adjustment, low-resolution simulation, gamma transformations, spatial transformations and mirroring.

Model training employed a composite loss function combining Dice loss and Cross-Entropy loss, integrated with deep supervision. Optimization was performed using Stochastic Gradient Descent (SGD) with an initial learning rate of 0.01 and a weight decay of 3 × 10^-5^. A polynomial learning rate scheduler was applied throughout training, which was conducted in a patch-wise manner with a patch size of [64 × 224 × 160] and a batch size of 2. The model was trained for 1000 epochs on an NVIDIA A40 GPU.

### 4) Data analysis postprocessing

Before calculating tortuosity, fractal dimension, and branch length, we applied several post-processing steps to clean the vasculature skeletons generated by ASN:

1. Skeleton Dilation: ML-generated vasculature skeletons can sometimes be fragmented, either due to missing vessel segments or minor discontinuities involving just a few voxels. To reconnect vessels split by small gaps, we applied a small 1-iteration binary dilation to the segmentation mask^42^.
2. Skeletonization: After dilation, we skeletonized^43, 44^ the segmentation mask to reduce vessel structures to a one-voxel-wide representation suitable for graph-based analysis.
3. Graph Construction: We constructed a bidirectional graph where each skeleton voxel was connected to its 26 possible neighbors. This enables straightforward detection of endpoints (nodes with one neighbor) and branch points (nodes with three or more neighbors). After graph construction, we retained only the largest connected component.
4. Branch Point and Endpoint Grouping: Due to imperfections in segmentation, particularly at bifurcations, multiple adjacent voxels could be incorrectly classified as separate branch points. To resolve this, we applied DBSCAN^45^ clustering to group nearby points and retained only the point closest to the cluster center.
5. Small Vessel Removal: Finally, we removed branches shorter than 9 voxels, as these were likely artifacts or incomplete vessel segments not fully captured by ASN.

### 5) Tortuosity estimation

There are various definitions for arterial tortuosity, some motivated by an easy manual estimation procedure for single or a small number of arteries, others more amenable for an automatic estimation^1, 2, 7, 16^. They all have in common that they intend to measure the same property, namely the elongation of arteries beyond the shortest possible path, and thus have been found to quite generally provide qualitatively similar information (but see Kashyap et al.^46^, Martelli & Giacomozzi^47^). Beyond the estimation formula, the definition of the vessel section onto which the estimation is being applied to is of importance. For example, from branchpoint to branchpoint (bifurcating), branchpoint or other seed point to endpoint (terminating), or along the whole vessel. The latter is difficult to implement, as vessels are branching, and the selection of the main vessel is somewhat arbitrary. Here we used the branch tortuosity also used in the BRAVA data set, defined by Diedrich et al.^48^, on the post-processed ASN labels.

## 3. Results

### 1) Validation of the ASN model

An example subject’s MRA, its ground truth COSTA labels, its ASN labels, and skeletonized labels are shown in Fig. 1. Visual inspection of the complete data set revealed the following. Overall, the ASN labels showed more vascularity than the ground truth data. However, not all of it reflected cerebral arteries; in other words, sporadic venous components or extracerebral blood vessel were visible. The ASN images sometimes showed more occipital arteries that were often not visible in the ground truth. Finally, the ASN model sometimes lacked or only had incomplete internal carotids at the base of the brain and sporadic lack of other vascular tree components.

**Figure 1:**
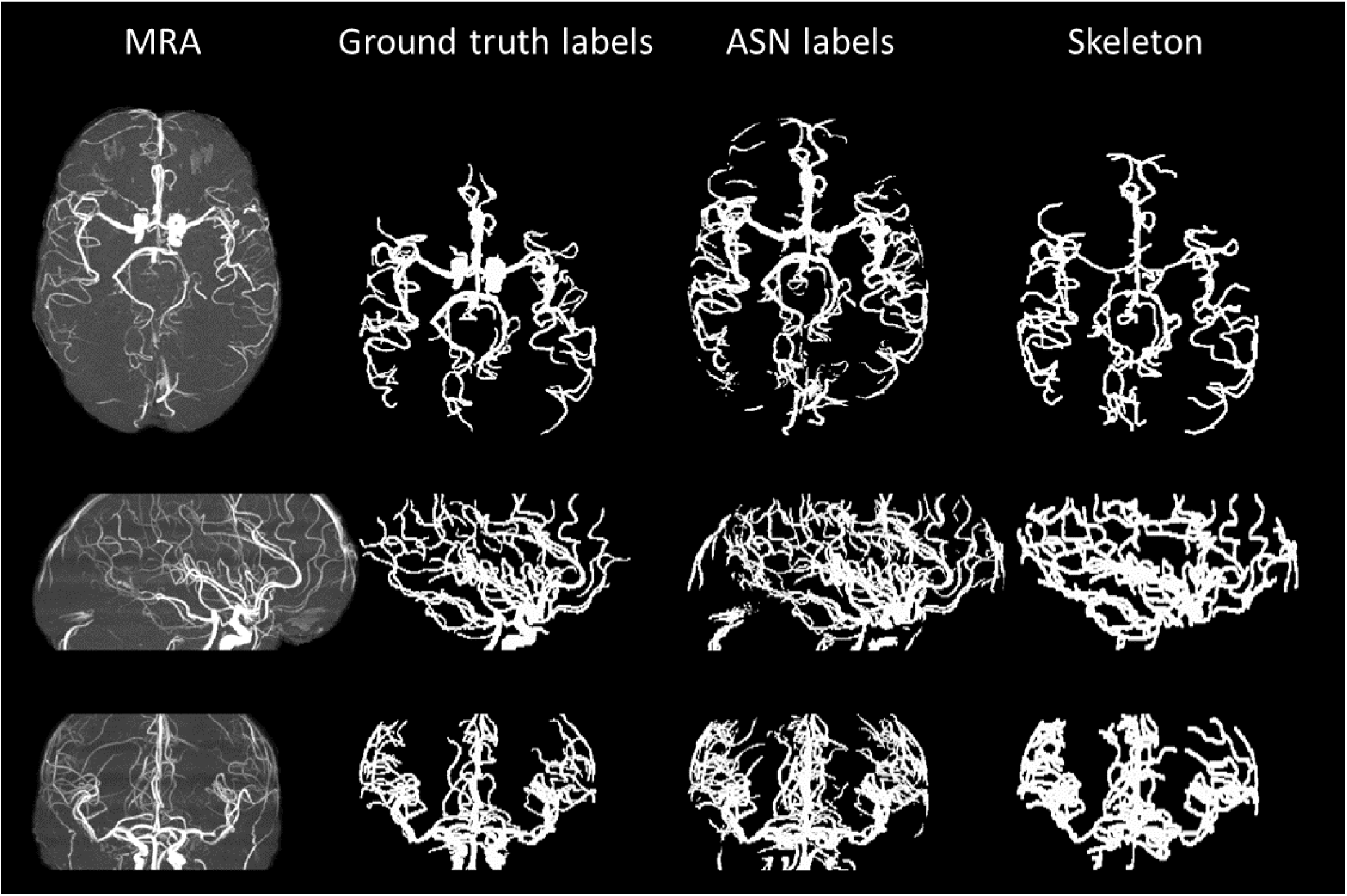
Comparison of ground truth COSTA labels and ASN labels for one representative subject. The first column shows MIPs of the MR arteriogram, the second column MIPs of the ground truth labels, the third column MIPs of the ASN labels, and the fourth column the skeleton derived from these labels. The skeleton follows from the first three post-processing steps in the Methods section and was dilated for visibility. The three rows correspond to axial, sagittal, and coronal views.

For a more quantitative analysis, an additional 61 cases from the COSTA dataset were used to evaluate the model’s performance using the Dice score. On this test set, the ASN model achieved a mean Dice score of 0.95 with a standard deviation of 0.02, and a median Dice score of 0.95.

### 2) Validation of the ASN model by tortuosity-age law

When the ASN model was applied to the unprocessed BRAVA MRA images (N = 56), the relationship between age and tortuosity could be modeled by a linear regression with R = 0.29 (p = 0.03), indicating a statistically significant increase of tortuosity with age (Fig. 2A). When terminating and bifurcating branches were analyzed separately, the correlation remained marginally significant for terminating branches (R = 0.28, p = 0.04), and only suggested a weak correlation for bifurcating branches (R = 0.19, p = 0.15) (Fig. 2B). For comparison, the semi-manually segmented MRA data reported by Wright et al.^8^ showed stronger correlations: R = 0.32 (p < 0.0006) for terminating branches and R = 0.44 (p < 0.0002) for bifurcating branches. Another summary marker, the mean fractal dimension, increased significantly with age (Fig. 2C; R = 0.37, p = 0.005). This finding again is in concordance with the results of Wright et. al, who also observed a significant increase of fractal dimension with age. The mean branch length did not change with age (Fig. 2D). However, and again in concordance with Wright et al., the bifurcating part of the branch length correlated with age (not shown; R = 0.27, p = 0.047)

**Figure 2:**
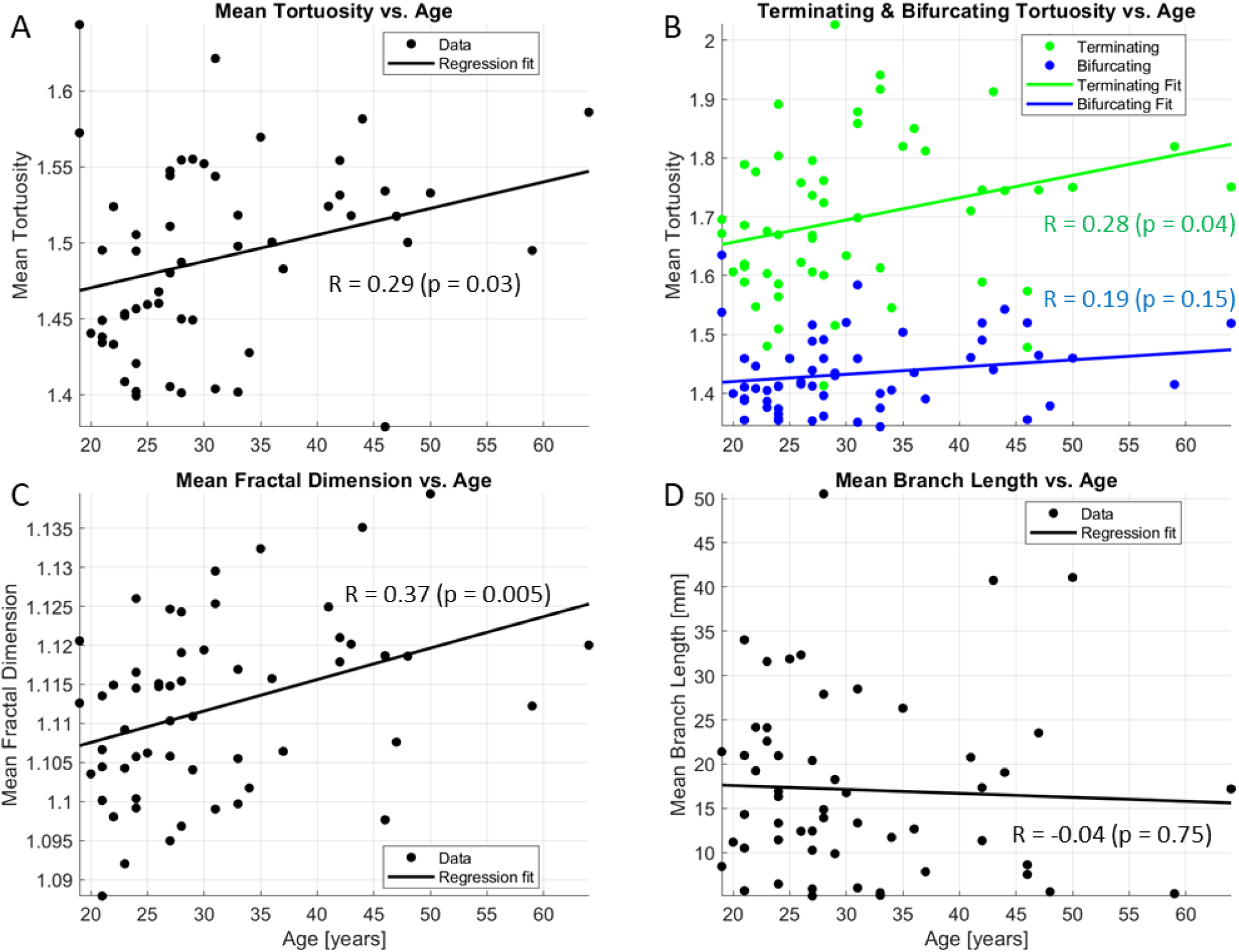
Tortuosity for the BRAVA data set in dependence of subject’s age (A) and a comparison of bifurcating vs. terminal branch estimations of tortuosity vs. age (B). Also, fractal dimension (C) and branch length (D) vs. age.

### 3) The age dependence of tortuosity in a large data set

Next, the BRAVA and IXI data sets were combined to a large data set to test the hypothesis of an age dependence of the tortuosity in a larger sample (N = 478). When the ASN model was applied to the unprocessed images of the combined BRAVA-IXI data set, the relationship between age and tortuosity could be modeled by a linear regression with R = 0.24 (p = 9 × 10^-8^), indicating a statistically highly significant increase of tortuosity with age (Fig. 3A). When terminating and bifurcating branches were analyzed separately, the correlation remained significant and positive both for terminating branches (R = 0.14, p = 0.002) and bifurcating branches (R = 0.25, p = 2 × 10^-8^) (Fig. 3B). The mean fractal dimension increased significantly with age (Fig. 3C; R = 0.23, p = 2 × 10^-7^). The fact that the mean branch length did not change with age (Fig. 3D; R = 0.02, p = 0.73) is notable and will be discussed later.

**Figure 3:**
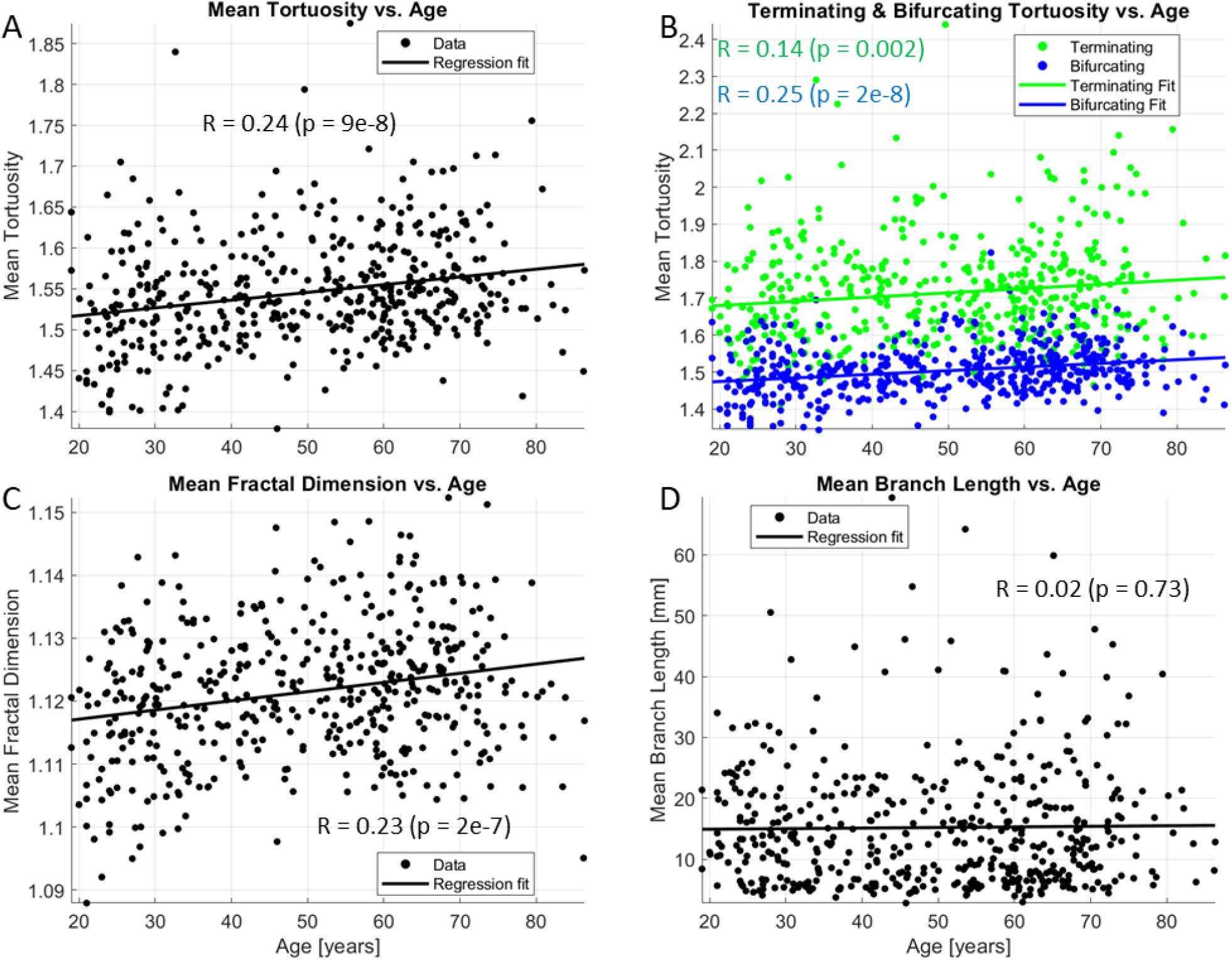
Tortuosity for the combined data set in dependence of subject’s age (A) and a comparison of bifurcating vs. terminal branch estimations of tortuosity vs. age (B). Also, fractal dimension (C) and branch length (D) vs. age.

## 4. Discussion

### 1) Validation of the ASN model by the ground truth

Whereas the ASN labels of cerebral arteries were less accurate than the semi-manually derived ground truth labels, the ASN labels still allowed for a useful rendering of cerebral arteries, with a high Dice similarity coefficient between ASN and ground truth COSTA labels. Notably, ASN is applicable to MRA data of varying image resolution and quality, including those typically encountered in clinical TOF-MRA. The observed tortuosity–age relationship was derived from datasets acquired at different resolutions, demonstrating the method’s robustness across standard imaging protocols. We expect that an application of the trained model to MRAs of different scanner manufacturers and imaging protocols would provide serviceable results.

### 2) Validation of the ASN model by tortuosity—age law

In the comparison of tortuosity from our ASN model segmentation with the tortuosity from the BRAVA data^8^, it turned out that our regression was less significant globally and with respect to the terminating branches, and not significant for the bifurcating branches. A weaker statistic was to be expected, as our fully automated segmentation is less accurate than a semi-manual segmentation of data. Importantly, the general positive correlation of tortuosity could be reproduced even in this relatively small data set.

### 3) The age dependence of tortuosity

We could reproduce the published finding of a positive correlation of cerebroarterial tortuosity in a large data set of MRAs with high significance (p = 9 × 10^-8^).

The fact that the mean branch length did not change with age (R = 0.02, p = 0.73; Fig. 3D) is interesting, as one would expect that with increasing tortuosity the branch lengths have to increase, too. A more detailed look into bifurcating vs. terminating branches shows that indeed the bifurcating branch lengths suggest an increase with age (R = 0.08, p = 0.07), whereas the terminating branch lengths decrease with age (R = −0.14, p = 0.002). Based on a review of scans from older patients, it seems that the model underperforms in these patients, capturing less information in terminating branches. The reasons for this underperformance might be the model itself, rooted in the physiology of aging, e.g. age-related changes in blood flow or vessel wall thickness, or even in the way the TOF-MRI captures aging blood vessels compared to younger blood vessels. For example, MRI parameters such as flip angles, echo times, and repeat times affect the MRA contrast^49-52^, and it might be necessary to actually adjust those to the age of the subjects in order to reduce bias. These and alternative hypotheses have been discussed previously^7^. To eliminate the possibility that the model itself falls short, one could use larger training data sets for older patients; only 15% of the subjects in the IXI-COSTA training dataset are 50 years or older.

The scale at which tortuosity was investigated was determined by the visibility of arteries in typical clinical MRAs. Using ultra-high field MRI and contrast agents, significantly smaller scales can be reached^10^. Delineating small arteries at these scales and distinguishing them from veins is a labor-intensive task. The authors hope that, in the future, machine learning approaches such as the one presented here may assist in this important challenge given the potential significance of small arteries to the health of the aging brain^53-56^. In a similar way, a possible dependence of arterial tortuosity on the brain region^6, 48, 57^ has not been investigated here but may be possible with machine learning in the foreseeable future, too.

## 5. Conclusion

The increase of arterial tortuosity with age in the human brain is a phenomenon that has been found in histopathological and imaging studies based on small to intermediate sized data sets. A crucial step in imaging studies is the segmentation of the arterial tree from the images, which are usually not perfect renderings of the vasculature. We are proposing a machine learning based approach for the segmentation step in magnetic resonance angiography images. Our ASN model has been trained and validated on a previously published ground truth data set. In addition, we could approximately reproduce the age dependence law of arterial tortuosity that was previously published in another data set, as well in an application to a large data set (N = 478). The trained ASN model can be used for an instant estimation of arterial tortuosity from individual MRA data, providing a summary marker for the subject’s cerebrovascular status.

## Data Availability

The ASN model for generating a brain arteriogram from data is available on GitHub: https://github.com/rachitsaluja/ASN

## Acknowledgements

The authors thank Armaan K. Tewary for help with manual segmentation of arteriograms, initial literature exploration, and early training and data preprocessing scripts.

## CRediT authorship contribution statement

Jorge T. Gomez: Conceptualization, Data curation, Validation, Writing – original draft, methodology, review & editing

Rachit Saluja: Conceptualization, Data curation, Validation, Writing – original draft, methodology, review & editing

Mert R. Sabuncu: Supervision, Writing – review & editing

Henning U. Voss: Supervision, Conceptualization, Writing – original draft, review & editing

## Declaration of generative AI and AI-assisted technologies in the writing process

During the preparation of this work the authors used the ChatGPT (May 2025 version) large language model (https://chat.openai.com/) in order to search for additional references and to improve the style of some paragraphs. After using this tool/service, the authors reviewed and edited the content as needed and take full responsibility for the content of the publication.

